# General purpose large language models match human performance on gastroenterology board exam self-assessments

**DOI:** 10.1101/2023.09.21.23295918

**Authors:** Shuhaib Ali, Omer Shahab, Reem Al Shabeeb, Farah Ladak, Jamie O. Yang, Girish Nadkarni, Juan Echavarria, Sumbal Babar, Aasma Shaukat, Ali Soroush, Bara El Kurdi

**Affiliations:** University of Texas Health, Department of Medicine, Division of Gastroenterology and Hepatology, San Antonio, Texas, USA; Division of Gastroenterology, Virginia Hospital Center, Arlington, Virginia, USA; Department of Medicine, Inova Fairfax Medical Campus, Falls Church, Virginia, USA; University of California Los Angeles, Department of Internal Medicine, Los Angeles, CA, USA; University of Texas Health, Department of Medicine, Division of Infectious Diseases, San Antonio, Texas, USA; Division of Gastroenterology, Department of Medicine, NYU Grossman School of Medicine, New York, NY, USA; Henry D. Janowitz Division of Gastroenterology, Icahn School of Medicine at Mount Sinai, New York, New York, USA; Division of Data-Driven and Digital Medicine (D3M), Icahn School of Medicine at Mount Sinai, New York, New York, USA; The Charles Bronfman Institute of Personalized Medicine, Icahn School of Medicine at Mount Sinai, New York, New York, USA

## Abstract

**Introduction:** While general-purpose large language models(LLMs) were able to pass USMLE-style examinations, their ability to perform in a specialized context, like gastroenterology, is unclear. In this study, we assessed the performance of three widely available LLMs: PaLM-2, GPT-3.5, and GPT-4 on the most recent ACG self-assessment(2022), utilizing both a basic and a prompt-engineered technique.

**Methods:** We interacted with the chat interfaces of PaLM-2, GPT-3.5, and GPT-4. We first applied a basic prompt approach, providing each exam question and answer text with minimalist text descriptions of any images. For the engineered approach, we added additional context and instructions. We assessed each model-prompt combination in terms of overall and difficulty-stratified performance and compared this to average human performance. We also evaluated each model’s self-assessed uncertainty. The highest scoring model-prompt combination was further assessed on the 2021 exam. We also assessed the impact of image descriptions on our findings.

**Results:** Using a basic prompt, PaLM-2, GPT-3.5, and GPT-4 achieved scores of 32.6%, 55.3%, and 68.9% respectively. With the engineered prompt, scores improved to 42.7%, 65.2%, and 76.3% respectively. Testing GPT-4 on the ACG-2021 exam yielded a similar score(75.3%). GPT-4 scores matched the average score for human test-takers reported by ACG(75.7%). GPT-4 showed a capability to self-assess its confidence accurately in the context of a multiple-choice exam with its confidence estimates falling within 5% of its actual performance. Excluding image-based questions didn’t change the primary findings.

**Discussion:** Our study highlights the capability of GPT-4 to answer subspecialty board-exam questions at a level commensurate with the average human test-taker. The results confirm that prompt-engineering can enhance LLMs’ performance on medical reasoning tasks. We also show GPT-4 can provide insightful measures of uncertainty in the setting of board-style multiple-choice questions, alerting users to low-quality answers. Future studies of LLMs in gastroenterology should incorporate prompt-engineering to maximize model capabilities.

**WHAT IS KNOWN:**

1. State of the Art large language models like GPT-4 and PaLM-Med 2 have achieved above average performance on USMLE board examinations.
2. In a previous study using basic model prompt instructions, GPT 3.5 and GPT 4 did not pass the 2021 and 2022 ACG self-assessment exams.

**WHAT IS NEW HERE:**

1. Optimizing large language model prompt instructions improved the performance of chat-based GPT-3.5, GPT-4, and PaLM 2 on the ACG self-assessment exams.
2. With optimized prompt instructions, chat-based GPT-4 performed at the level of average human test takers on ACG-self assessment examinations and achieved a passing score.
3. Chat-based GPT-4 self-reported confidence levels correlated with correct answer rates on the ACG-self assessment examinations.

## Introduction

State of the art large language models (LLMs) have become increasingly capable of a diverse range of tasks, ranging from natural language processing to predicting protein sequences. These LLMs are utilize the transformer deep learning architecture (1) and self-supervised learning (2,3) to train on massive datasets. This results in a foundational artificial intelligence mode that is capable of a broad range of tasks without significant additional model training. The general-purpose capabilities of these LLMs has already begun to have an impact across various industries, such as medicine (4–6).

Previous generation LLMs have already been tailored for medical use cases such as PubMedBERT (7), ClinicalBERT(8) with limited success. Another model; GatorTron (9) shortly followed, it was trained on PubMed, Wikipedia, and EHR clinical note text, but was only able to perform a limited set of natural processing tasks. The first LLM to achieve a passing score on USMLE-style questions, Med-PaLM, is an instruction-tuned version of Google’s Flan-PaLM model (10) that is further “prompt tuned” to align model responses to the requirements of the medical domain (11). Newer LLMs such as the next-generation Med-PaLM 2 (12) as well as OpenAI’s GPT-3.5(13) and GPT-4(14) follow the same general instruction-based framework of the Flan-PaLM model, but with even larger training datasets and further model alignment (12). With these advances, the most advanced GPT-4 and Med-PaLM2 models have achieved expert-level performance on most standardized medical reasoning metrics with only a minimal degree of prompt engineering (12,15). Both successfully and comfortably passed the USMLE examination with a cushion of more than 20% (12,15).

Though the models performed well with USMLE board exam questions, their performance on specialty and subspecialty board exams that are less likely to be present in the training data and require an even higher level of expertise is still a subject of debate. A growing number of studies have assessed this in different medical specialties including radiology (16), otolaryngology (17), neurosurgery (18), ophthalmology (19) and(20), orthopedic surgery (21), and bariatric surgery (22) (**Supplementary Table 1**). A recent study evaluated baseline GPT-3.5 and GPT-4 gastroenterology reasoning performance using the 2021 and 2022 American College of Gastroenterology’s Self-Assessment exams (ACG-SA), finding that both models did not score high enough to pass (23). Notably, this study did not employ the prompt engineering methods utilized in key prior medical reasoning papers, which have previously been shown to improve LLM reasoning performance (24–26). Surprisingly, GPT-3.5 outscored the newer GPT-4 in this study, in sharp contrast to prior studies. Additionally, the study did not compare LLM performance to human test takers.

To interrogate the gastroenterology reasoning capabilities of current generation LLMs further, we explored the effect of prompt engineering and assessed LLM performance versus humans at the global and question levels. In this study, we assessed the performance of GPT-3.5 and GPT-4 and the now publicly available PaLM 2.

## Methods

### Exam Question Pre-Processing

Because access to image-capable LLMs has been limited, previous studies have excluded questions with images to isolate the models’ text-based reasoning capabilities (12,15,16). Since our question set was small and contained a large proportion of questions with images, we elected 1) to assess if a provided image was necessary to answer the associated question correctly and 2) to provide text descriptions for any necessary images. Two gastroenterologists individually performed the evaluations and generated text descriptions that conveyed descriptive visual information without providing any high-level classification or key buzzwords which could reveal the answer independently. For example, an image of a perianal abscess was described as “a small perianal bulge with overlying redness and surrounding erythema.” Discrepancies in evaluations and descriptions were resolved by consensus.

### Large Language Model Approach

For this study, all three language models were accessed in May 2023, directly through their respective public chat interfaces: Bard (PaLM 2), ChatGPT (GPT-3.5), and ChatGPT plus (GPT-4). The 2022 ACG-SA examination was first provided to all three LLMs (PaLM 2, GPT-3.5, and GPT-4) to determine the best performing model. Each question was entered in succession following the same order provided by the test. A new chat session was started for each unique exam, model, and prompt strategy combination.

To assess baseline model capabilities, we elicited exam answers using a basic prompt strategy of only providing each individual question and its answer choices. To assess more advanced latent reasoning capabilities, we applied a more advanced engineered prompt strategy. For this approach, the following additional text was added before the basic prompt: “You are a gastroenterology fellow undergoing board examination. You will be given a list of multiple-choice questions. Please read each question carefully and assess/consider each choice separately step by step before providing your answer. You should also state your degree of confidence in your answers on a scale from 0 to 100. Do this systematically and in a step-by-step fashion.”

In the prompt engineered approach, including additional contextual information allows the LLM to better calibrate its responses (26). Asking the model to think step-by-step elicits the “chain-of-thought” method of augmenting model reasoning by forcing the model to generate descriptive text in a step-by-step manner (25). Finally, LLM self-confidence assessments in the context of multiple-choice questions have been shown to have reasonable accuracy (27–29).

The LLM and prompting strategy that collectively demonstrated the best performance was additionally validated on the ACG-SA 2021 exam.

### Performance Metrics

Answer choices generated by each LLM were compared against the exam answer key to calculate the overall, and difficulty-stratified percentage of correct answers for each LLM. Average scores for human test takers were kindly provided by ACG upon request. To evaluate performance based on questions difficulty, we stratified the questions into 4 categories based on percentage of human respondents that answered them correctly: 12-40% was considered very challenging, 40-60% was considered challenging, 60-80% was considered moderate and 80-100% was considered easy. To generate a calibration curve for each model, exam questions were grouped based on the model’s confidence in its answer into 4 categories, mean model confidence for each category was plotted against the mean actual percentage correct. A calibration plot was used to represent this as described by Nori et al. (15). To assess the impact of our image-based descriptions on our analysis, we used a two-sample t-test with a significance level of 0.05 to compare LLM performance on questions with and without images.

## Results

### Exam Characteristics

The 2022 ACG-SA examination consisted of 300 questions, of which 137 contained images (**Supplementary Table 2**). For 36 of these image-based questions, the image was either not required to answer the question or the question text provided an adequate description. The 2021 ACG-SA examination contained the same total number of questions, but contained 114 with images. Of those image-based questions, only seven could be answered without supplemental information. Neither of the assessed examinations is publicly available and accessible for inclusion in LLM training data.

### LLM Performance

Using the basic prompting strategy, PaLM 2 achieved a score of 32.6% correct answers, while GPT-3.5 and GPT-4 scored 55.3% and 68.9% respectively (**Table 1**). PaLM 2 was unable to answer 76 questions, citing its limited capability to provide clinical responses (e.g., “I’m a text-based AI, and that is outside of my capabilities”). When these questions were excluded, PaLM 2’s performance improved to 43.8%. Using the engineered prompt strategy, the scores of PaLM 2, GPT-3.5, and GPT-4 improved to 42.7%, 63.0%, and 76.3% respectively. PaLM 2 could not provide answers to only 26 questions with this approach, which was improved compared to the basic prompting strategy (when these questions were excluded, performance improved to 47.8%).

**Table 1:**
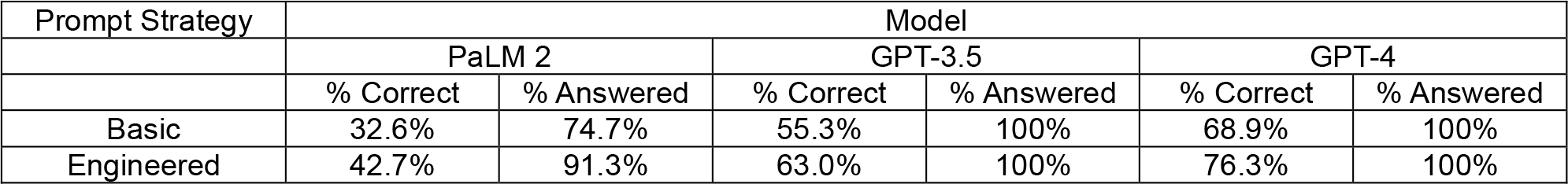
Overall LLM Performance on ACG-SA 2022.

Using an engineered prompt strategy with GPT-4 produced the highest exam score. To confirm the validity of these results, we applied this approach to the 2021 ACG-SA examination. The model performance remained consistent, scoring 75.3% on the older exam. When adjusting for question difficulty, GPT-4’s performance based on question difficulty was consistently within the range of human performers for both the 2021 and 2022 exams, while GPT-3.5’s lagged closely behind, and PaLM-2 significantly underperformed at all levels. Data shared by ACG showed average performance by human test takers at 75.7%(**Table 2**).

**Table 2:** LLM Performance, Stratified by Question Difficulty.

|  | Humans<br>(% correct) | GPT-4 2022<br>(% correct) | GPT-4 2021<br>(% correct) | GPT-3.5 2022<br>(% correct) | PaLM-2 2022<br>(% correct) |
| --- | --- | --- | --- | --- | --- |
| All Questions | 75.7% | 76.3% | 75.3% | 63.0% | 42.7% |
| Question difficulty |  |  |  |  |  |
| Very Challenging | 12-40% | 26.9% | 31.6% | 26.9% | 11.5% |
| Challenging | 40-60% | 70.2% | 59.1% | 51.4% | 32.4% |
| Medium | 60-80% | 71.1% | 73.1% | 56.7% | 40.0% |
| Easy | 80-100% | 89.1% | 87.5% | 61.9% | 54.4% |

### Uncertainty Assessment

The engineered prompt strategy also sought to self-assess model confidence for each exam answer. PaLM 2 was unable to perform this task. GPT-4 self-characterized its response certainty between 40% and 95% and generally matched its actual aggregated performance within a range of 5%. In contrast, GPT 3.5 self-assessed its certainty between 40% and 95%, but its aggregate response accuracy never surpassed 75%. At its most confident, GPT-3.5 overestimated its actual performance by more than 20%. (**Table 3, Figure 2**).

**Table 3:** LLM performance based on self-reported uncertainty (predicted probability that a provided answer is correct).

| Mean predicted probability | GPT-4 |  | GPT-3.5 |  | PaLM 2 |  |
| --- | --- | --- | --- | --- | --- | --- |
|  | Questions (n) | Correct (%) | Questions (n) | Correct (%) | Questions (n) | Correct (%) |
| 55% | 53 | 56.6% | 45 | 51.1% | N/A | N/A |
| 75% | 46 | 67.4% | 93 | 54.8% | N/A | N/A |
| 85% | 148 | 79.7% | 140 | 70.7% | N/A | N/A |
| 95% | 53 | 88.7% | 22 | 72.7% | N/A | N/A |

**Figure 1.**
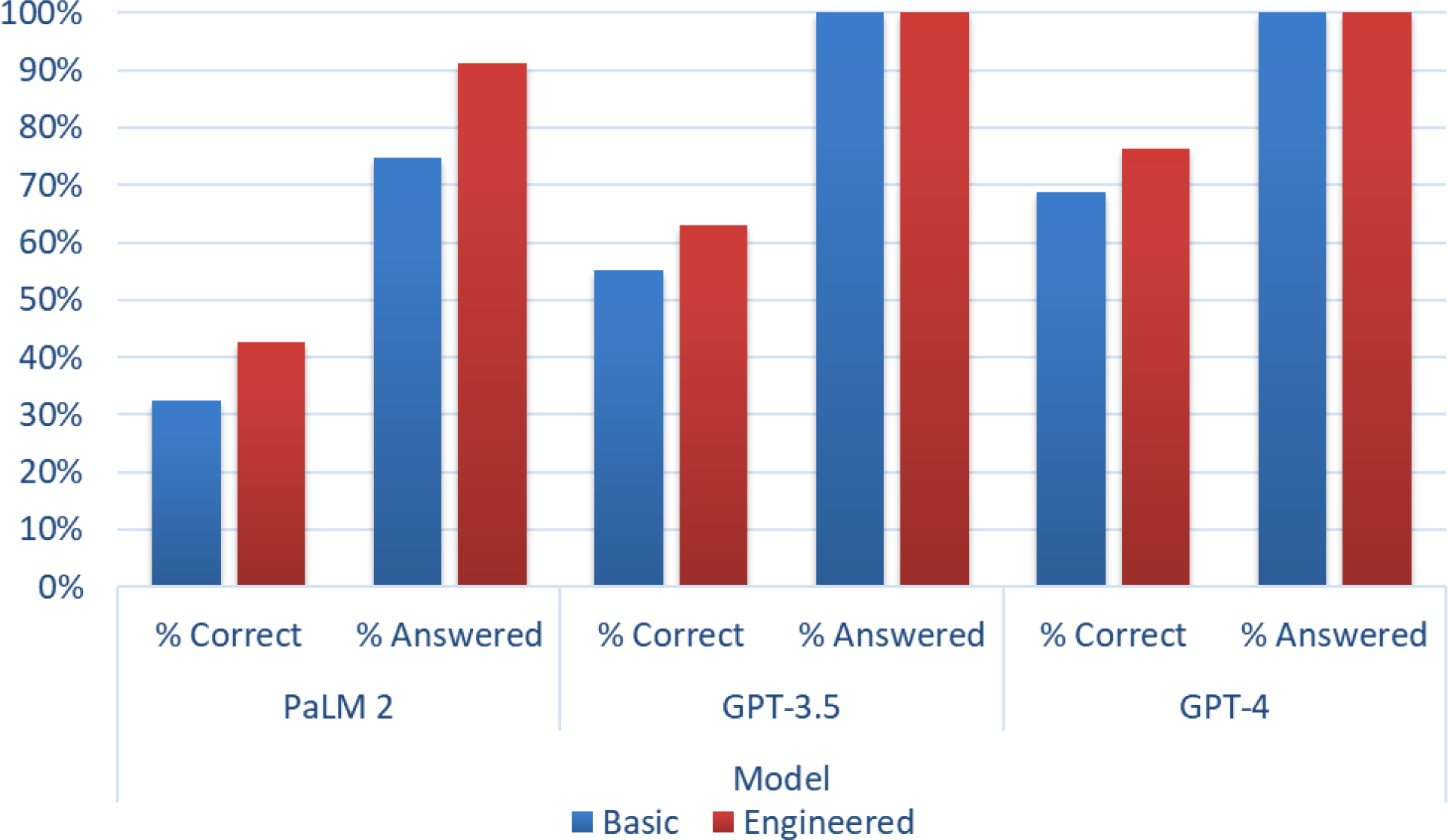
LLM performance on ACG-SA 2022 stratified by model and approach.

**Figure 2:**
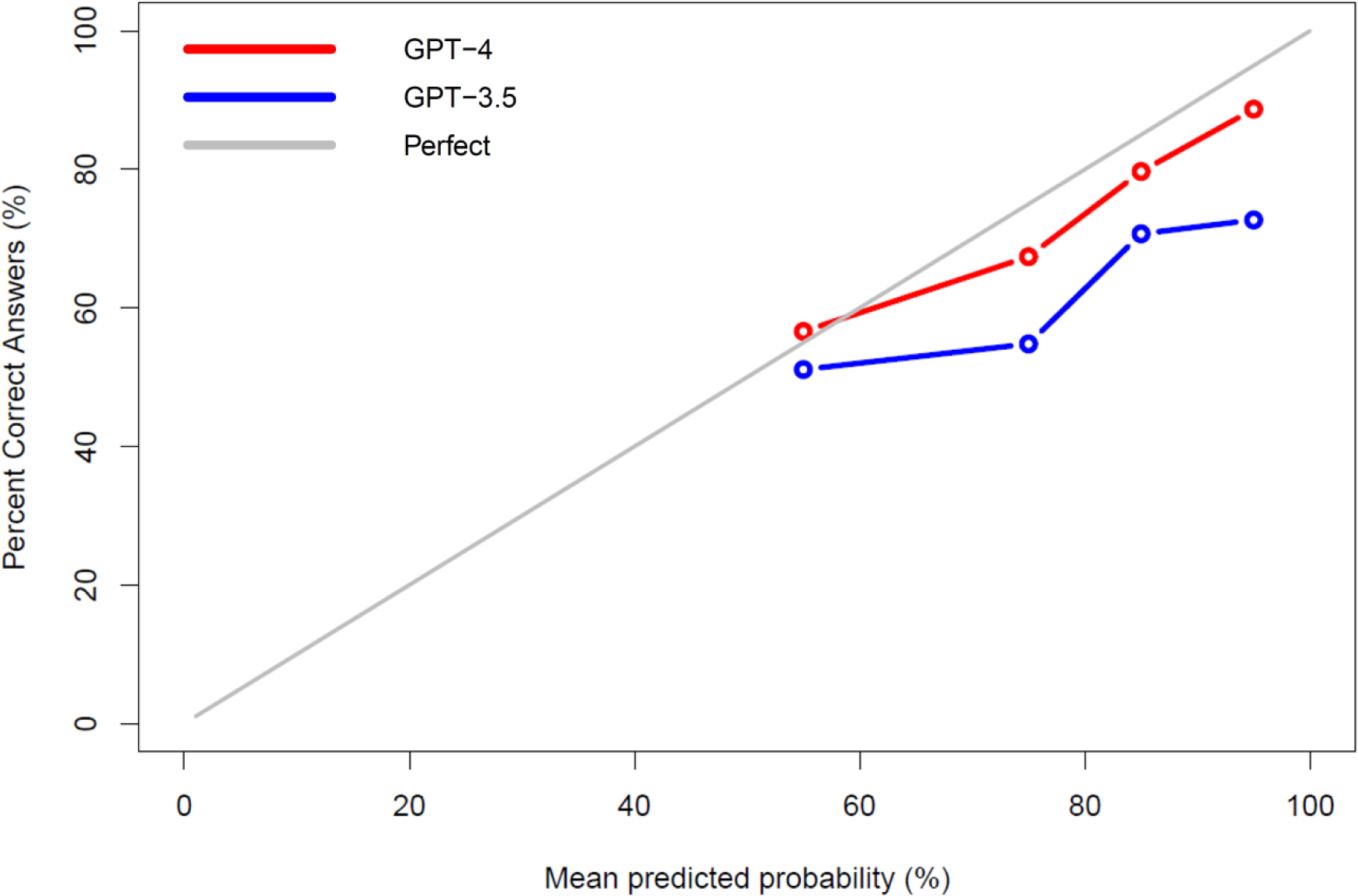
Large Language Model Response Confidence Calibration Curve

### Image Description Sensitivity Analysis

No significant difference in performance was observed across models for questions with or without images (Table 2). This applied to both basic and engineered approaches.

## Discussion

In this study, we found that even a simple prompt engineering strategy can significantly enhance the performance of LLMs on subspecialty clinical examinations. With prompt engineering, GPT-4 obtained passing scores on both the 2021 and 2022 ACG-SA exams and performed consistently within the same range as human test takers overall as well as for every question difficulty level. We also found that self-reported uncertainty measurements were reasonably accurate for GPT-4, suggesting that prompts that include uncertainty measures may help alert users to low quality responses.

While GPT-3.5’s performance improved with the engineered prompt, it failed to reach the passing threshold of 70%. Additionally, it significantly overestimated its true response accuracy by significant margins at the upper ends of self-reported confidence, indicating that its uncertainty measures are unreliable when the model is reporting high levels of confidence. These findings highlight the significant improvements in newer iterations of the GPT model and parallel previously published studies(15).

In contrast, we found that the base PaLM 2 model is poorly suited for use in the medical domain. PaLM 2 repeatedly declined to provide answers to the exam questions and refused to perform any self-assessments of certainty, likely due to model limitations imposed at the system level. However, PaLM 2 had much worse performance than both GPT-3.5 or GPT-4, even when excluding refused questions. It is likely that the medically tailored Med-PaLM 2 model performs better with the type of assessment in this study, but at the time of publication we did not have access to this more tailored model.

No meaningful difference in our results were noted when excluding image questions with our supplementary image descriptions, indicating that this approach can be used to increase dataset pool until future models are able to process medical images.

Our findings contrast with those of a highly publicized recent study by Suchman et al.(23), which showed that GPT-3.5 and GPT-4 could not pass the 2021 and 2022 ACG-SA exams. We suspect there are several reasons for the differences between the results of our studies. The differences in performance when using a basic prompt strategy may be due to underlying improvements in the GPT models or perhaps subtle differences in how each study interfaced with the LLM. Our largest performance gain, however, was from the use of a prompt engineering strategy incorporating additional contextual information and a chain of thought instruction.

The training data and model architecture of GPT-4 are not known, making it difficult to understand how or why GPT-4 is able to reason to such a high degree on medical board exams. It is likely that the model was trained, at minimum, on all available open-source text datasets including textbooks, Wikipedia, PubMed abstracts, and open-access journal articles. Like all LLMs, GPT-4 “reasons” by using a statistical model to serially predict chunks of text, or “tokens”(14). Prompt engineering methods help guide LLMs to prioritize text responses that are well-reasoned and correct and to narrow the knowledge sources used for reasoning tasks(30). LLM-type reasoning appears to be well-suited for multiple-choice question reasoning, but it is unclear if this extends to the more ambiguous reasoning of day-to-day clinical tasks.

Our study has several limitations. Our engineered prompt approach did not include more advanced prompt engineering such as few-shot prompting, self-consistency, ensemble refinement, or response chaining methods to enhance reasoning performance (12). We elected to avoid these to demonstrate the benefit of even a simple, no-code prompt-based approach.

We also avoided changes to model temperature and system prompts. These approaches could improve outcomes but require software expertise that most clinicians do not possess and therefore, maybe out of reach for our intended audience. Additionally, the ACG self-assessment is an imperfect exam and does not reflect all aspects of medical reasoning used in routine clinical practice. Finally, our human-generated image descriptions may be biasing model performance for image-based questions and would not reflect a real-world use case for the LLMs. It is notable that the tested LLMs consistently performed slightly better on questions without images, indicating that image descriptions may be useful but do not replace the need for independent image analysis.

Despite these limitations, our study has several strengths. We have the broadest range of data assessing LLMs in the gastroenterology space to date, assessing the impact of simple prompt engineering, comparing model performance to human test takers, and examining self-assessment capabilities for all three widely available chat-based LLMs. Our assessment of a no-code prompt engineering approach allows the average clinician to understand and implement our methodology for future research. We also describe a new approach for inclusion of image-based questions in test datasets, increasing the pool of questions to be considered in future LLM research.

The high level of subspeciality board exam medical reasoning demonstrated by GPT-4 with a simple prompt engineering strategy holds great promise for gastroenterology medical education and other clinical reasoning tasks. Self-paced chatbot-based tutors the Khan Academy’s Khanmigo (31) could be adapted for subspecialty medical training, increasing access to expert clinical knowledge and reasoning and democratizing medical education. Future work should assess multimodal models that can ingest multiple data types (e.g., text and images) as well as the integration and application of additional reasoning augmentation strategies.

With further refinements and appropriate validation of medical reasoning capabilities, state of the art LLMs could also power intelligent clinical decision support tools, clinical workflows, and patient-support interfaces. LLM augmentation techniques such as model finetuning (32), advanced prompt engineering (26), prompt chaining (33), and database linkage (34) have been applied successfully to improve reasoning in other contexts. Smaller, more efficient models are also being developed (35), which will improve model speed and reduce computational costs. Before integrating LLMs into clinical practice, a comprehensive characterization of their clinical reasoning capabilities with real-world clinical scenarios is needed.

## Conclusion

We find that GPT-4 was able to pass the ACG self-assessment exam with an engineered prompt, performing within the range of average human test takers and providing insightful measures of uncertainty. While other models failed this examination, the success of GPT-4 suggests that further applications in clinical education and practice may become possible as these models and our skills in interfacing them continue to evolve.

## Data Availability

All data produced in the present study are available upon reasonable request to the authors.

---

**Supplementary Table 1:**
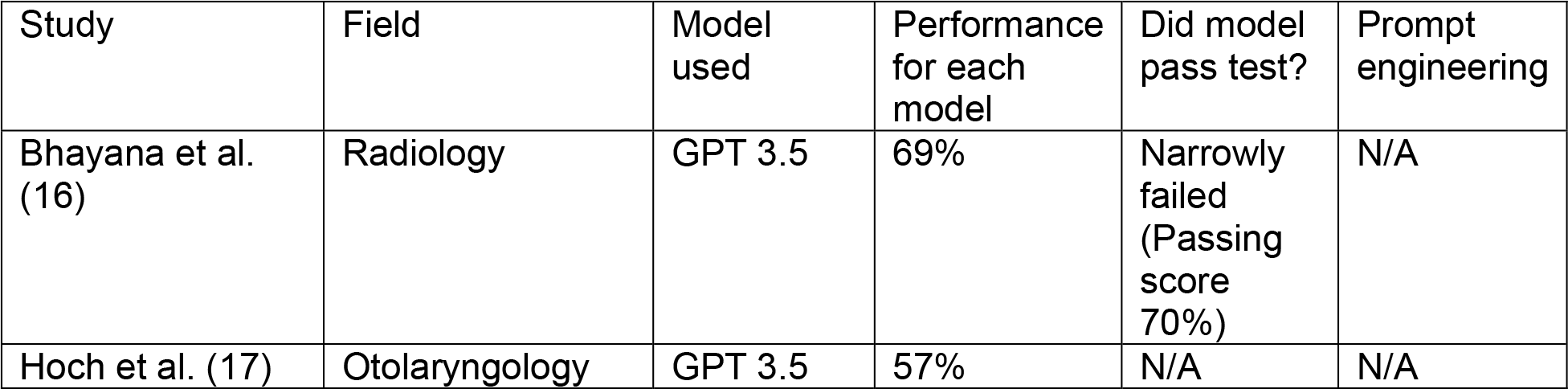
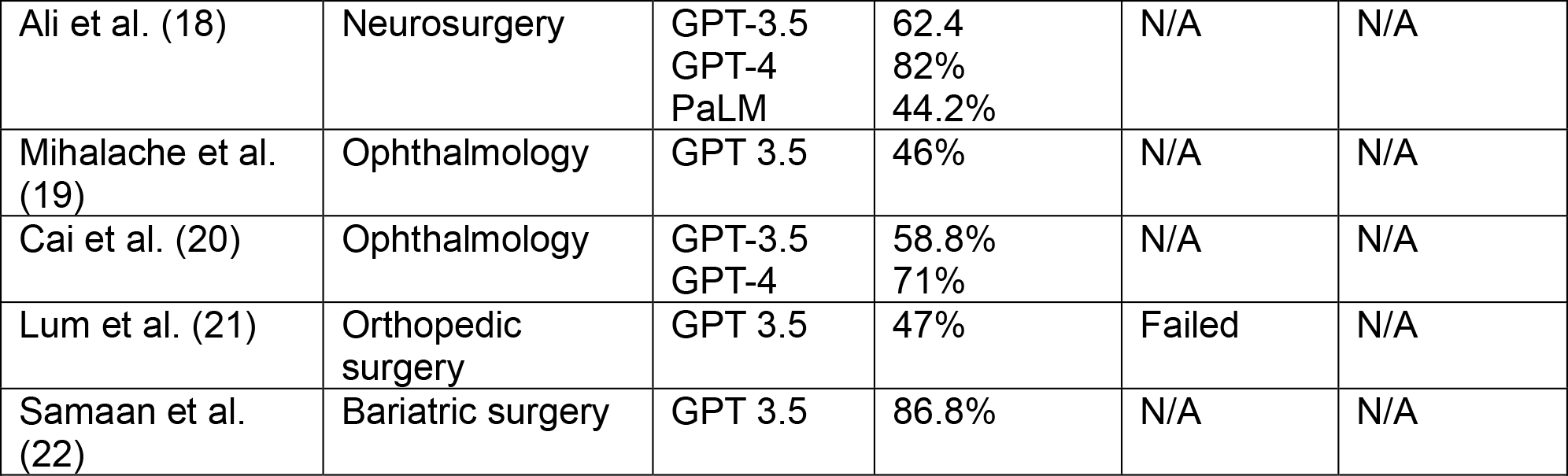
LLM performance on specialty and subspecialty board exams. (N/A = not applicable or not reported).

**Supplementary Table 2:**
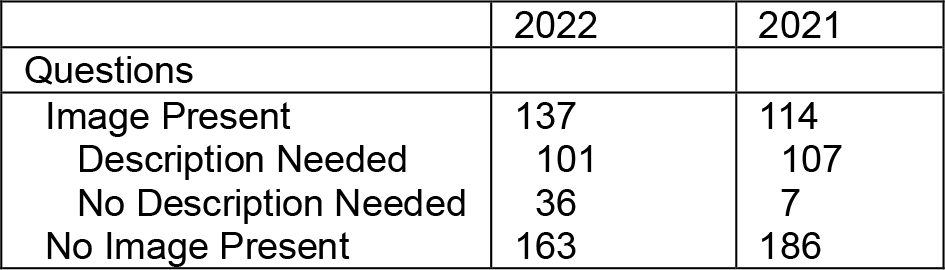
Exam Characteristics.

|  | 2022 | 2021 |
| --- | --- | --- |
| Questions |  |  |
| Image Present | 137 | 114 |
| Description Needed | 101 | 107 |
| No Description Needed | 36 | 7 |
| No Image Present | 163 | 186 |

**Supplementary Table 2:** The impact of image descriptions on model performance (2022 ACG-SA).

|  | PaLM 2 |  |  | GPT-3.5 |  |  | GPT-4 |  |  |
| --- | --- | --- | --- | --- | --- | --- | --- | --- | --- |
|  | No image questions (% correct) | With image descriptions (% correct) | p-Value | No image questions (% correct) | With image descriptions (% correct) | p-Value | No image questions (% correct) | With image descriptions (% correct) | p-Value |
| Basic | 27.8% | 32.6% | 0.27 | 56.9% | 55.3% | 0.31 | 69.7% | 68.2% | 0.50 |
| Engineered | 43.0% | 42.7% | 0.87 | 63.2% | 63.0% | 0.97 | 73.5% | 75.7% | 0.46 |

## Notes

### Competing Interest Statement

The authors have declared no competing interest.

### Funding Statement

This study did not receive any funding

